# Presentation collateral patterns and outcomes after endovascular thrombectomy for large vessel occlusion stroke

**DOI:** 10.1101/2022.01.29.22270093

**Authors:** Robert W Regenhardt, Michael H Lev, Julian He, Adam A Dmytriw, Justin E Vranic, James D Rabinov, Christopher J Stapleton, Aman B Patel, Aneesh B Singhal, R Gilberto Gonzalez

## Abstract

**Background:** Endovascular thrombectomy (EVT) revolutionized large vessel occlusion (LVO) stroke. However, treatment decisions and prognostication are challenging without advanced imaging. We sought to determine the relationship of simple CTA collateral patterns and outcomes after EVT.

**Methods:** We identified patients with anterior LVO who underwent guideline based EVT from a single center from 2019-2020. Inclusion criteria were available CTA and 90-day modified Rankin Scale (mRS). Arterial-phase CTA collaterals were categorized as malignant, other, or symmetric.

**Results:** Among 74 patients, the median age was 75 and 49% were female. Collaterals were symmetric (36%), malignant (24%), or other (39%). Comparing collateral patterns there were no differences in demographics, presentations, time from last well, good reperfusion, or intracerebral hemorrhage. Median NIHSS was 18 for malignant, 19 for other, and 11 for symmetric (p=0.02). Intracranial ICA occlusions were present in 28% of malignant, 3% of other, and 11% of symmetric (p=0.04). Ninety-day mRS ≤2 was achieved in 17% of malignant, 38% of other, and 67% of symmetric. Collateral pattern was a significant determinant of 90-day mRS ≤2 (aOR=6.62, 95%CI=2.24,19.53; p=0.001) in a multivariable model including age, NIHSS, baseline mRS, thrombolysis, LVO location, and good reperfusion.

**Conclusions:** Simple CTA collateral pattern is a robust determinant of 90-day outcomes after EVT. Further prospective studies are needed to understand how collateral pattern can guide EVT treatment decisions and long-term prognosis.

## Introduction

Large vessel occlusion (LVO) stroke accounts for the largest proportion of stroke-related death and disability.^1^ Acute care for patients with LVO stroke has been revolutionized by endovascular thrombectomy (EVT).^2–4^ However, treatment selection and prognostication can be challenging when advanced imaging, such as CT perfusion (CTP) or MRI, is unavailable.^5^ Unfortunately, community hospitals and underserved regions are often without these resources.^6,7^ This is especially true for patients in the extended window or with unknown onset.^8^ Furthermore, the transfer of patients to thrombectomy capable centers can be delayed, which reduces the likelihood of EVT.^9^

Collaterals are alternative vessels, consisting of primary circle of Willis and secondary pial-pial leptomeningeal anastomoses, that can compensate for reduced blood flow in the setting of LVO.^1^ Collateral patterns vary dramatically among patients with stroke and are highly related to infarct growth.^10^ Indeed, the collateral pattern assessed by presentation CT angiography (CTA) may be an appropriate proxy for infarct volume and infarct growth rate.^11^ In patients with LVO not treated with reperfusion therapies, symmetric collateral pattern on CTA had a sensitivity of 87% and a specificity of 94% for 24-hour infarct volume <50 cc.^12^ We sought to determine the relationship of presentation simple CTA collateral patterns and outcomes after EVT.

## Results

Among 74 patients who met inclusion criteria from 2019 to 2020, the median age was 75 (IQR 58-82), and 49% were female. Collaterals were symmetric (36%), malignant (24%), or other (39%). Comparing collateral patterns there were no differences in demographics, risk factors, time from last known well, thrombolysis treatment, TICI 2b-3 reperfusion, or sICH. Median NIHSS was 18 (IQR 14-23) for malignant, 19 (IQR 12-22) for other, and 11 (IQR 8-18) for symmetric (p=0.02). Intracranial ICA occlusions were present in 28% of malignant, 3% of other, and 11% of symmetric (p=0.04) (Table 1).

**Table 1.** Patient demographics, risk factors, presentations, treatments, and outcomes.

|  | Total | Malignant | Other | Symmetric | P-Value |
| --- | --- | --- | --- | --- | --- |
| Age, Median (IQR) | 75 (58,82) | 71 (52,82) | 76 (59,81) | 72 (59,81) | 0.72 |
| Female | 36 (49%) | 7 (39%) | 16 (55%) | 13 (48%) | 0.55 |
| Black | 4 (5%) | 2 (11%) | 1 (3%) | 1 (4%) | 0.47 |
| White | 49 (66%) | 11 (61%) | 16 (55%) | 22 (82%) | 0.10 |
| Asian | 5 (7%) | 1 (6%) | 3 (10%) | 1 (4%) | 0.60 |
| Hispanic | 3 (4%) | 1 (6%) | 1 (3%) | 1 (4%) | 0.93 |
| Baseline mRS $\geq 3$ | 5 (7%) | 2 (11%) | 2 (7%) | 1 (4%) | 0.62 |
| Atrial Fibrillation | 21 (28%) | 5 (28%) | 8 (28%) | 8 (30%) | 0.98 |
| Coronary Disease | 23 (31%) | 7 (39%) | 11 (38%) | 5 (19%) | 0.21 |
| Diabetes Mellitus | 19 (26%) | 3 (17%) | 9 (31%) | 7 (26%) | 0.55 |
| Dyslipidemia | 30 (41%) | 6 (33%) | 11 (38%) | 13 (48%) | 0.57 |
| Heart Failure | 14 (19%) | 4 (22%) | 7 (24%) | 3 (11%) | 0.42 |
| Hypertension | 52 (70%) | 13 (72%) | 18 (62%) | 21 (78%) | 0.43 |
| Obesity/Overweight | 31 (42%) | 6 (33%) | 14 (48%) | 11 (41%) | 0.59 |
| Previous Stroke | 11 (15%) | 3 (17%) | 2 (7%) | 6 (22%) | 0.27 |
| Previous TIA | 4 (5%) | 1 (6%) | 1 (3%) | 2 (7%) | 0.81 |
| Chronic Renal Insufficiency | 9 (12%) | 2 (11%) | 3 (10%) | 4 (15%) | 0.87 |
| Smoker | 9 (12%) | 1 (6%) | 6 (21%) | 2 (7%) | 0.19 |
| Cardioembolism | 42 (57%) | 9 (50%) | 18 (62%) | 15 (56%) | 0.71 |
| Large Artery Atherosclerosis | 10 (14%) | 3 (17%) | 2 (7%) | 5 (19%) | 0.40 |
| Cryptogenic | 14 (19%) | 3 (17%) | 4 (14%) | 7 (26%) | 0.49 |
| Dissection | 2 (3%) | 2 (11%) | 0 (0%) | 0 (0%) | 0.04 |
| Hypercoagulability | 6 (8%) | 1 (6%) | 5 (17%) | 0 (0%) | 0.06 |
| NIHSS, Median (IQR) | 17 (10,22) | 18 (14,23) | 19 (12,22) | 11 (8,18) | 0.02 |
| Intravenous Thrombolysis | 21 (28%) | 6 (33%) | 11 (38%) | 4 (15%) | 0.14 |
| Telestroke Consult | 35 (47%) | 6 (33%) | 12 (41%) | 17 (63%) | 0.11 |
| Transfer | 43 (58%) | 8 (44%) | 15 (52%) | 20 (74%) | 0.10 |
| LKW-Telestroke | 1.73 (1.10,4.17) | 1.38 (0.99,1.73) | 1.53 (1.10,3.25) | 3.27 (1.23,6.00) | 0.18 |
| LKW-Hub Arrival | 3.08 (0.90,5.07) | 2.59 (1.81,3.25) | 2.63 (0.67,4.72) | 4.47 (2.24,6.68) | 0.14 |
| LKW-Arterial Puncture | 3.60 (2.70,6.07) | 3.42 (2.82,4.22) | 3.03 (2.22,5.37) | 5.25 (2.72,7.79) | 0.16 |
| Intracranial ICA Occlusion | 9 (12%) | 5 (28%) | 1 (3%) | 3 (11%) | 0.04 |
| M1 MCA Occlusion | 51 (69%) | 13 (72%) | 23 (79%) | 15 (56%) | 0.15 |
| M2 MCA Occlusion | 14 (19%) | 0 (0%) | 5 (17%) | 9 (33%) | 0.02 |
| Stent Retriever EVT | 26 (37%) | 8 (44%) | 10 (39%) | 8 (31%) | 0.64 |
| Aspiration Only EVT | 44 (63%) | 10 (56%) | 16 (62%) | 18 (69%) | 0.64 |
| TICI 2b-3 | 65 (88%) | 15 (83%) | 26 (90%) | 24 (89%) | 0.80 |
| sICH | 5 (7%) | 3 (17%) | 1 (3%) | 1 (4%) | 0.16 |
| Discharge mRS $\leq 2$ | 13 (18%) | 1 (6%) | 3 (10%) | 9 (33%) | 0.02 |
| 90-Day mRS $\leq 2$ | 32 (43%) | 3 (17%) | 11 (38%) | 18 (67%) | 0.003 |

Abbreviations: IQR Interquartile Range, mRS modified Rankin Scale, TIA Transient Ischemic Attack, NIHSS National Institutes of Health Stroke Scale, LKW Last Known Well, ICA Internal Carotid Artery, MCA Middle
Cerebral Artery, EVT Endovascular Thrombectomy, TICI Thrombolysis in Cerebral Infarction, sICH symptomatic Intracerebral Hemorrhage.

Ninety-day mRS ≤2 was achieved in 17% of malignant, 38% of other, and 67% of symmetric (p=0.003) (Table 1, Figure 1). Collateral pattern was a significant determinant of 90-day mRS ≤2 (aOR=6.62, 95%CI=2.24,19.53; p=0.001) in a multivariable model including age (aOR=0.92, 95%CI=0.87,0.97; p=0.001), NIHSS (aOR=0.98, 95%CI=0.89,1.08; p=0.68), baseline mRS ≥3 (aOR=6.14, 95%CI=0.60,63.12; p=0.13), intravenous thrombolysis (aOR=2.14, 95%CI=0.52,8.91; p=0.29), occlusion location (aOR=0.53, 95%CI=0.16,1.82; p=0.31), and TICI 2b-3 reperfusion (aOR=10.45, 95%CI=1.05,104.3; p=0.05) (Table 2).

**Figure 1.**
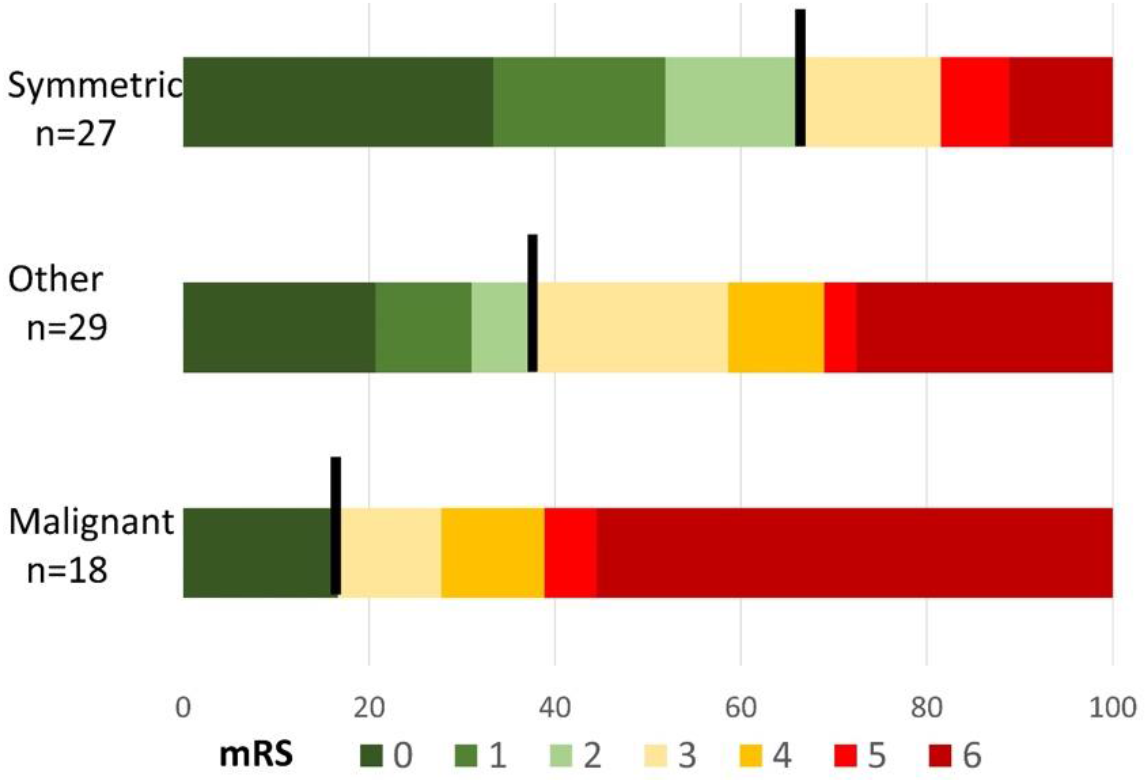
Ninety-day modified Rankin Scale (mRS) score comparing collateral patterns.

**Table 2.**
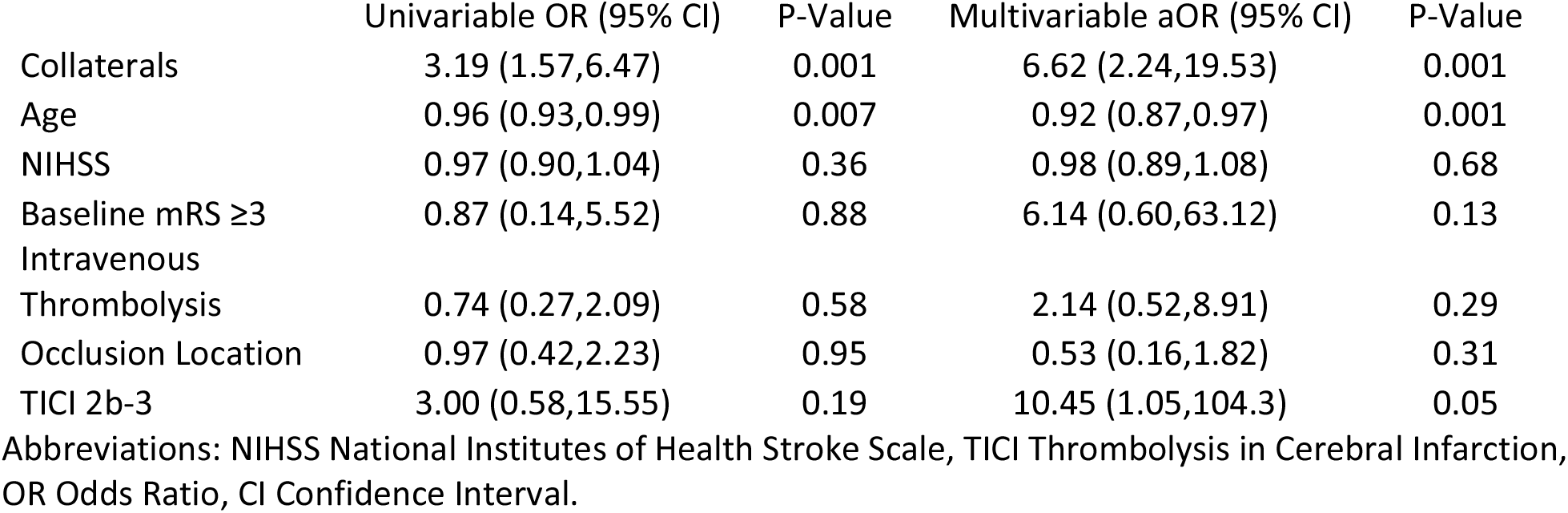
Determinants of 90-day modified Rankin Scale (mRS) score ≤2.

|  | Univariable OR (95% CI) | P-Value | Multivariable aOR (95% CI) | P-Value |
| --- | --- | --- | --- | --- |
| Collaterals | 3.19 (1.57,6.47) | 0.001 | 6.62 (2.24,19.53) | 0.001 |
| Age | 0.96 (0.93,0.99) | 0.007 | 0.92 (0.87,0.97) | 0.001 |
| NIHSS | 0.97 (0.90,1.04) | 0.36 | 0.98 (0.89,1.08) | 0.68 |
| Baseline mRS $\geq 3$ | 0.87 (0.14,5.52) | 0.88 | 6.14 (0.60,63.12) | 0.13 |
| Intravenous Thrombolysis | 0.74 (0.27,2.09) | 0.58 | 2.14 (0.52,8.91) | 0.29 |
| Occlusion Location | 0.97 (0.42,2.23) | 0.95 | 0.53 (0.16,1.82) | 0.31 |
| TICI 2b-3 | 3.00 (0.58,15.55) | 0.19 | 10.45 (1.05,104.3) | 0.05 |

Abbreviations: NIHSS National Institutes of Health Stroke Scale, TICI Thrombolysis in Cerebral Infarction, OR Odds Ratio, CI Confidence Interval.

## Discussion

In this retrospective analysis of patients who underwent EVT, there was significant variability in collateral patterns, with only 36% being symmetric. Symmetric collaterals were associated with less severe stroke presentation and good 90-day functional outcome. Even when controlling for age, stroke severity, baseline disability, thrombolysis treatment, occlusion location, and adequate reperfusion, collateral pattern was a significant determinant of 90-day outcome.

Among patients treated with EVT, we noted significant variability in collateral patterns. They were symmetric in 36%, malignant in 24%, and other in 39%. This is consistent with our local EVT treatment selection guidelines, which do not exclude patients based on collateral patterns. Indeed, others have reported a range of collateral quality, albeit assessed differently, among patients treated with EVT.^13,14^ In addition, there was a trend for malignant patterns to have shorter times from LKW, but this difference did not reach statistical significance. This may be related to our treatment exclusion of patients with poor ASPECTS or large infarcts since patients with poor collaterals have faster infarct growth.^12^ Furthermore, more proximal occlusion locations were more likely to have poor collaterals; ICA occlusions were present in 28% of malignant, 3% of other, and 11% of symmetric. This stands to reason, and early studies have explored the relationship between occlusion location and collateral quality.^15^

While there have been some prior analyses of the relationship of collaterals with stroke severity and 90-day outcomes, the data are mixed. Furthermore, we contend our assessment of collateral patterns has significant advantages compared to other approaches.^12^ Among our cohort, the median NIHSS was 18 for malignant patterns, 19 for other, and 11 for symmetric. Other studies corroborate our results, showing that patients with poor collaterals have higher NIHSS.^11,16^ Furthermore, 90-day mRS ≤2 was achieved in 17% of malignant, 38% of other, and 67% of symmetric within our cohort. This relationship persisted even when controlling for age, stroke severity, baseline disability, thrombolysis, occlusion location, and adequate reperfusion. There is mixed prior literature regarding this relationship, highlighting the need for further research.^11,16,17^

The mechanism of the relationship between collaterals and clinical outcomes may be explained by presentation infarct volume and infarct growth. Indeed, infarct volume is one of the strongest determinants of 90-day outcomes, even among patients who undergo EVT.^18^ In our prior study, worse collaterals were an independent determinant of both greater presentation infarct volume and infarct growth rate, even when controlling for other variables including occlusion site, NIHSS, and age.^12^ Beyond presentation, continued infarct growth may be important for outcomes even among those who achieve successful reperfusion with EVT.^19,20^ We previously demonstrated that collateral patterns continue to affect infarct growth up to 48 hours in patients not treated with EVT, suggesting there is no “collapse” of collaterals over this time.^21^

Importantly, there are several implications from this study. Our classification approach for collaterals has been previously described,^12^ and is consistent with other studies using a three-category approach.^22^ While CTP can be used to estimate collaterals,^10,22^ CTA is more widely available, has no threshold dependence, and allows direct visualization of the cerebral vessels. Our simple classification system is robust and immediately translatable. There is growing data that extended window EVT is beneficial even without advanced imaging.^23^ A simple non-contrast CT and single arterial phase CTA may be sufficient to triage most patients for EVT. Understanding collaterals may also aid in decision making for the inter-hospital transfer of patients to EVT-capable centers.^24,25^ At many smaller hospitals, MRI or CTP are not readily available.^6,26^

There are several limitations to consider. First, patients were identified, and collateral patterns were evaluated retrospectively. There may be some selection bias for patients treated with endovascular thrombectomy. Second, the single center design could limit generalizability. Third, the “other” collateral pattern category was heterogenous, with some patients rated as closer to symmetric and others rated as closer to malignant. Despite these potential limitations, this simple three-category classification system is easy to learn, easy to use, robust, and immediately translatable.

In conclusion, collateral pattern is a robust determinant of 90-day outcomes after thrombectomy. Further prospective studies are needed to evaluate the role collateral pattern in EVT treatment decisions and prognostication.

## Methods

This study was approved by the local institutional review board. Informed consent was waived based on minimal patient risk and practical inability to perform the study without the waiver. Patients who underwent EVT for anterior circulation LVO were identified retrospectively from a prospectively maintained database at a single referral center.^27^ Inclusion criteria were available CTA for retrospective review and prospectively recorded 90-day modified Rankin Scale (mRS) score.

CTA was performed using multi-detector scanners (GE Medical Systems) from the vertex to the aortic arch following injection of 65–140 ml of a nonionic contrast agent (Isovue; Bracco Diagnostics) at a rate of 3 to 4 ml/s. The median parameters were 1.25-mm slice thickness, 220 mm reconstruction diameter, 120 kV, and 657 mA. CTDI_vol_ ranged from 65-95 mGY, and DLP ranged from 2593-3784 mGY-cm.

Collateral patterns were interpreted by CAQ-certified neuroradiologists with over 25 years’ experience interpreting acute stroke studies (RGG, MHL).Interpreters were blinded to clinical presentations, treatments, other imaging, and clinical outcomes. Patterns were determined by visual review of the maximum intensity projection arterial phase CTA images, which were classified as “symmetric”, “malignant”, or “other”.^12^ Briefly, a symmetric pattern was defined as contrast visualized with similar or near similar conspicuity of the ischemic compared to the contralateral non-ischemic MCA territory. A malignant pattern was defined as no contrast visualized over at least 50% of the MCA territory at risk. “Other” was defined as any additional pattern, rated as intermediate between symmetric and malignant. Vessel occlusion site on CTA was documented as internal carotid artery (ICA) terminus, first (M1) middle cerebral artery (MCA) segment, and second (M2) MCA segment.^28^ Cervical ICA stenosis was defined as >70% by NASCET criteria.^29^

The local database included demographic information, medical history, clinical presentation, treatments, and outcomes for consecutive patients treated with EVT. Stroke severity (National Institutes of Health Stroke Scale, NIHSS) was determined as described.^30^ Alteplase treatment decisions were guideline-based at the discretion of a vascular neurologist. EVT treatment decisions were guideline-based at the discretion of the treating vascular neurologist and neurointerventionalist. Thrombolysis in cerebral infarction (TICI) scores were determined by a neurointerventionalist using the modified scale: 2a partial filling <50%, 2b partial filling ≥50%, 3 complete perfusion.^3^ Adequate reperfusion was defined as TICI 2b-3.^31^ Symptomatic intracerebral hemorrhage (sICH) was defined as any PH1 or PH2 by ECASS criteria associated with new symptoms during the hospitalization.^32^ 90-day modified Rankin Scale (mRS) was obtained by telephone call or follow-up clinic visit.^33,34^ Good functional outcome was defined as 90-day mRS ≤2.^35^

Median and interquartile range (IQR) were reported for continuous nonparametric variables. Percent and count were reported for categorical variables. Differences among three groups of nonparametric continuous variables were assessed using the Kruskal Wallis test. Associations with good functional outcome were assessed by logistic regression. Variables of interest were selected *a priori* for their possible relevance to good functional outcome. Distributions were assumed nonparametric based on the Kolmogorov-Smirnov and Shapiro-Wilk tests. Two-tailed P values <0.05 were considered statistically significant. Analyses were performed with Prism version 6.01 (GraphPad) and SPSS version 23.0 (IBM Corp).

## Data Availability

The data supporting these findings will be made available upon reasonable request and pending approval of the local institutional review board.

## Acknowledgments

Ms. Joyce A McIntrye maintained the local stroke database.

## Author Contributions

RWR, MHL, ABS, and RGG conceived the idea. MHL and RGG assessed collateral patters. ABS led clinical data collection. RWR, JDR, CJS, ABP performed thrombectomies. RWR, JH, and RGG led data analysis and interpretation. RWR prepared the first draft of the manuscript. All authors read, provided significant edits, and approved the final manuscript.

